# Serum DNA methylome of the colorectal cancer serrated pathway

**DOI:** 10.1101/2022.03.25.22272961

**Authors:** María Gallardo-Gómez, Lara Costas-Ríos, Carlos A. Garcia-Prieto, Lara Álvarez-Rodríguez, Luis Bujanda, Maialen Barrero, Antoni Castells, Francesc Balaguer, Rodrigo Jover, Manel Esteller, Antoni Tardío Baiges, Joaquín González-Carreró Fojón, Joaquín Cubiella, Loretta De Chiara

**Author notes:** Corresponding author: Loretta De Chiara, Department of Biochemistry, Genetics and Immunology, CINBIO, University of Vigo. Campus As Lagoas-Marcosende s/n. 36310 Vigo, Spain.

## Abstract

**Objective:** The clinical relevance of the serrated pathway of colorectal carcinogenesis is evident but the screening of serrated lesions remains challenging. We aimed to characterize the serum methylome of the serrated pathway, and to evaluate circulating cell-free DNA (cfDNA) methylation as a source of biomarkers for the non-invasive screening and diagnosis of serrated lesions.

**Design:** We collected serum samples from individuals with serrated adenocarcinoma (SAC), traditional serrated adenomas, sessile serrated lesions, hyperplastic polyps and with no colorectal findings. First, epigenome-wide methylation was quantified in cfDNA pooled samples with the MethylationEPIC array. Then, methylation profiles were compared to tissue and serum cfDNA datasets. Finally, biomarker utility of serum cfDNA methylation was evaluated.

**Results:** We identified a differential methylation profile that can distinguish high-risk serrated lesions from the absence of serrated neoplasia, showing concordance with tissue methylation from SAC and sessile serrated lesions (external datasets). We report that the methylation profiles in serum cfDNA are pathway-specific, clearly separating serrated lesions from conventional adenomas. Among the differentially methylated regions (DMR) we report, the combination of two DMRs within the genes *NINJ2* and *ERICH1* discriminated high-risk serrated lesions and SAC with 91.4% sensitivity and 64.4% specificity, while methylation from a DMR within *ZNF718* reported 100% sensitivity for the detection of SAC at 96% specificity.

**Conclusion:** This is the only study available to date exploring the serum methylome of serrated lesions. We have identified a differential methylation profile in serum specific to the serrated pathway. The serum methylome may serve as a source of non-invasive biomarkers for screening and detection of high-risk serrated lesions.

## INTRODUCTION

Colorectal cancer (CRC) is a very heterogeneous disease that develops via the stepwise accumulation of multiple genetic and epigenetic alterations.[1] While two-thirds of CRC arise from the classical adenoma-to-carcinoma sequence, also known as the chromosome instability (CIN) pathway, the serrated pathway accounts for 15-30% of CRC. The clinical and biological relevance of this alternative pathway has been pointed out in the last years [2–4]. Serrated lesions are a heterogeneous group of saw-toothed lesions, characterized histologically by a stellate pattern of crypt in-folding.[1,5] The most recent classification of the World Health Organization further defines the serrated lesions into four main categories, namely, hyperplastic polyps (HP), sessile serrated lesions (SSL, with and without dysplasia), unclassified serrated adenomas/polyps (SP), and traditional serrated adenomas (TSA).[5,6] HP are the most-frequently occurring serrated lesions (60-75%) and rarely undergo malignant transformation.[7] SSL account for 20-35% of all serrated lesions; dysplastic SSL are associated with advanced age, female sex, and proximal colon location.[7,8] TSA are the rarest form of serrated lesion (1-7%), found predominantly in the distal colon. Overall, TSA have a protuberant growth pattern with villiform projections, challenging the distinction between TSA and classical tubulovillous adenomas.[7,9]

A two-arm model has been proposed to describe the progression of the serrated pathway, characterized by genetic and epigenetic alterations that initiate and drive malignant transformation. The V600E activating *BRAF* mutation is a distinguishing trait of the serrated pathway and one of its first detected events, present in 70-80% of HP, >90% of SSL, and 20-40% of TSA.[4,10] After initiating *BRAF* mutations, serrated tumors may develop via two different routes: (i) one converging with the MSI pathway, characterized by mutations in the DNA mismatch machinery repair or by *MLH1* hypermethylation, leading to SSL that may progress to tumors with MSI-high phenotype; alternatively, (ii) lesions with *BRAF* mutations can acquire *TP53* mutations, activating oncogenic signaling such as Wnt and TGF-β, and epithelial-to-mesenchymal transition, resulting in TSA and microsatellite stable (MSS) tumors.[1,11] Tumors developing by either arm usually present high levels of CpG island methylation. The CpG island methylation phenotype (CIMP) is characterized by the hypermethylation of gene promoter regions causing loss of tumor suppressor gene function and can be already detected at early tumor stages.[12,13] CIMP status correlates with poorly differentiated tumors containing *BRAF* mutations and MSI, commonly located in the proximal colon, and mostly in older female patients.[12,14]

Detection and removal of premalignant lesions is one of the principal objectives of CRC screening. Large or dysplastic SSL and TSA are considered the precursor lesions of the serrated pathway.[9,15] While TSA are similar to conventional adenomas regarding the development of cancer, the subset of serrated lesions acquiring MSI, such as SSL, have an accelerated transition to carcinomas (1-3 years).[11] This, together with the fact that SSL are flat, with a mucus cap and indistinct borders that make them likely to be missed and incompletely resected during colonoscopy, suggests that a large proportion of interval CRC, those developed within the recommended surveillance window of 3-5 years, arise from the serrated pathway.[5,8,16] The similarities in histological appearance between SSL and HP also result in misclassification and misdiagnosis.[5]

Also, SSL are less prone to bleed due to their subtle morphology and thick but diffuse vascularity; thus, the fecal immunochemical test (FIT) commonly used for CRC screening has limited sensitivity for the detection of serrated lesions regardless of size or presence of dysplasia.[17–19] Hence, the successful screening of serrated lesions is still unmet.

In this study, we performed an epigenome-wide analysis of serum cfDNA pools to explore the methylation signatures in patients with serrated lesions, with the aim to characterize the serum methylome as a potential source of non-invasive biomarkers that could be implemented in screening programs to improve the rate of detection of serrated lesions and reduce the incidence of serrated CRC.

## RESULTS

### Study overview

This study was conducted to explore the serum methylome of precancerous lesions belonging to the serrated pathway of colorectal carcinogenesis in a prospective multicentre cohort. Individuals were grouped into five main categories: (i) serrated adenocarcinoma (SAC), (ii) high-risk serrated polyps (HR-SP) comprising traditional serrated adenomas (TSA), sessile serrated lesions (SSL), and serrated polyps (SP) with dysplasia or ≥ 10 mm; (iii) high-risk hyperplastic polyps (HR-HP), defined as HP ≥ 10 mm; (iv) low-risk serrated lesions (LR-SL) including SP without dysplasia < 10 mm and HP < 10 mm; and (v) healthy individuals with no colorectal findings (NCF). First, epigenome-wide methylation levels were quantified in pooled cfDNA samples to characterize the differential methylation profile between no serrated neoplasia (NSN: NCF and LR-SL) and high-risk serrated lesions (HR-SL: HR-HP and HR-SP); concordance with tissue methylation levels was assessed using external datasets. Then, the pathway-specific cfDNA methylation signature was evaluated together with cfDNA pools from the conventional CRC pathway. Finally, targeted assays were performed to evaluate the potential biomarker utility of serum cfDNA methylation to detect serrated lesions in an independent cohort. Clinical and epidemiological characteristics of patients included in the study are summarized in Table 1.

**Table 1.** Clinical and epidemiological characteristics of patients included in the study. The number of patients, age median and range, sex, and colorectal findings is provided. CRC was staged according to the American Joint Committee on Cancer (AJCC) system. CRC: colorectal cancer, FFPE: Formalin-fixed paraffin-embedded, HGD: high-grade dysplasia, HP: hyperplastic polyp, HR-HP: high-risk hyperplastic polyp, HR-SL: high-risk serrated lesion, HR-SP: high-risk serrated polyp, LGD: low-grade dysplasia, LR-SL: low-risk serrated lesion, MS-qPCR: methylation-specific quantitative PCR, NCF: no colorrectal findings, SAC: serrated adenocarcinoma, SP: serrated polyp; SSL: sessile serrated lesion; TSA: traditional serrated adenoma. ^†^SAC tissue samples are paired with a healthy mucosa sample from the same patient.

|  |  | Epigenome-wide analysis<br>(n=106, serum) |  | Biomarker evaluation<br>by MS-qPCR<br>(n=80, serum) |  | Matched<br>tumor-mucosa<br>(n=6, FFPE tissue) |
| --- | --- | --- | --- | --- | --- | --- |
|  |  | HR-SL | NSN | HR-SL | NSN | SAC<sup>†</sup> |
| **Total (n)** |  | **46** | **60** | **35** | **45** | **6** |
| **Age median**<br>**(range)** |  | 62.0<br>(51-73) | 61.5<br>(51-74) | 64.0<br>(52-73) | 60.0<br>(52-76) | 73.5<br>(63-90) |
| **Sex** |  |  |  |  |  |  |
|  | Male | 23 | 30 | 16 | 25 | 2 |
|  | Female | 23 | 30 | 19 | 20 | 4 |
| **NCF** |  | - | **30** | - | **20** | - |
| **LR-SL** |  | - | **30** | - | **25** | - |
|  | HP < 10mm | - | 27 | - | 18 | - |
|  | SP < 10mm | - | 0 | - | 3 | - |
|  | SSL < 10mm | - | 3 | - | 4 | - |
| **HR-HP (HP > 10mm)** |  | **16** | - | **10** | - | - |
|  | Distal location | 8 | - | 5 | - | - |
|  | Proximal location | 8 | - | 5 | - | - |
| **HR-SP** |  | **30** | - | **20** | - | - |
|  | TSA | 4 | - | 4 | - | - |
|  | Large/dysplastic SSL | 14 | - | 13 | - | - |
|  | SP > 10mm | 26 | - | 12 | - | - |
|  | HGD | 13 | - | 9 | - | - |
|  | LGD | 2 | - | 8 | - | - |
|  | Distal location | 15 | - | 8 | - | - |
|  | Proximal location | 15 | - | 12 | - | - |
| **SAC** |  | - | - | **5** | - | **6** |
|  | Stage I | - | - | 2 | - | 1 |
|  | Stage II | - | - | 0 | - | 1 |
|  | Stage III | - | - | 2 | - | 4 |
|  | Stage IV | - | - | 1 | - | 0 |
|  | Distal location | - | - | 2 | - | 3 |
|  | Proximal location | - | - | 3 | - | 3 |

### cfDNA sample pooling

For the serum methylome profiling, 106 serum cfDNA samples (Table 1) were grouped into 11 cfDNA pools as described in.[20] Briefly, five men and five women with the same colorectal pathology were included in each pool, matched by recruitment hospital and age (median 62, range 51-74). A detailed description of the pooled samples can be found in Supplementary Table 1. No pools were assembled for SAC cases due to limited sample availability. The final quantity of cfDNA in the pools ranged from 124 to 336 ng. There was no statistically significant difference in the mean age between pools (ANOVA, *p*-value < 0.05).

### Serum methylome profiling of the serrated pathway

Differential methylation analyses were performed across the total of 734,739 CpG-targeting probes left after quality filtering and normalization. Pairwise comparisons were carried out between the four pathological groups (NCF, LR-SL, HR-HP, and HR-SP) according to the expected progression of the serrated pathway: either NCF - LR-SL - HR-HP, or NCF - LR-SL - HR-SP. Following this scheme, six independent differential methylation analyses were performed: NCF vs LR-SL, NCF vs HR-HP, NCF vs HR-SP, LR-SL vs HR-HP, LR-SL vs HR-SP, and HR-HP vs HR-SP. Differential methylation results are detailed in Supplementary Figures 1-3.

To explore the differential methylation profile of precursor serrated lesions, pooled samples were grouped into no serrated neoplasia (NSN: NCF and LR-SL) and high-risk serrated lesions (HR-SL: HR-HP and HR-SP). After the comparison between HR-SL and NSN, only one CpG site achieved a significant false discovery rate (FDR, Bonferroni-Hochberg correction) lower than 10%, likely due to the small group size available. This CpG site was hypomethylated in HR-SL with a methylation difference of 14.66%. Thus, to explore serum methylome profiles we used a less stringent statistical criterion: CpG sites with a nominal *p*-value < 0.01 and at least 10% difference in methylation levels were considered as differentially methylated positions (DMPs). Applying this criteria, we identified 330 DMPs between HR-SL and NSN (Supplementary Table 2). Among them, 30.3% (100/330) were hypermethylated, while 69.7% (230/330) were hypomethylated in HR-SL (Figure 1A). One of the hypermethylated DMPs, cg24917382, is located within a CpG island of *IGF2* promoter, gene which is part of one of the proposed panels to classify CIMP tumors.[21] The methylation levels for each DMP and the sample clustering profile are shown in the heatmap in Figure 1B. Regarding functional genomic elements, 39.1% of DMPs were related to CpG islands (CGI, CGI-shores, and CGI-shelves) and 26.7% were associated to gene promoter regions including TSS200, TSS1500 (200 bp and 200-1,500 bp upstream of the transcription starting site, respectively), 5’UTR, and first exons (Figure 1C). Hypermethylated DMPs were enriched in intergenic regions, while hypomethylated DMPs were enriched in intergenic regions and CGI-shelves (one-sided Fisher’s exact test odds-ratio > 1 and *p*-value < 0.05) (Figure 1D).

**Figure 1.**
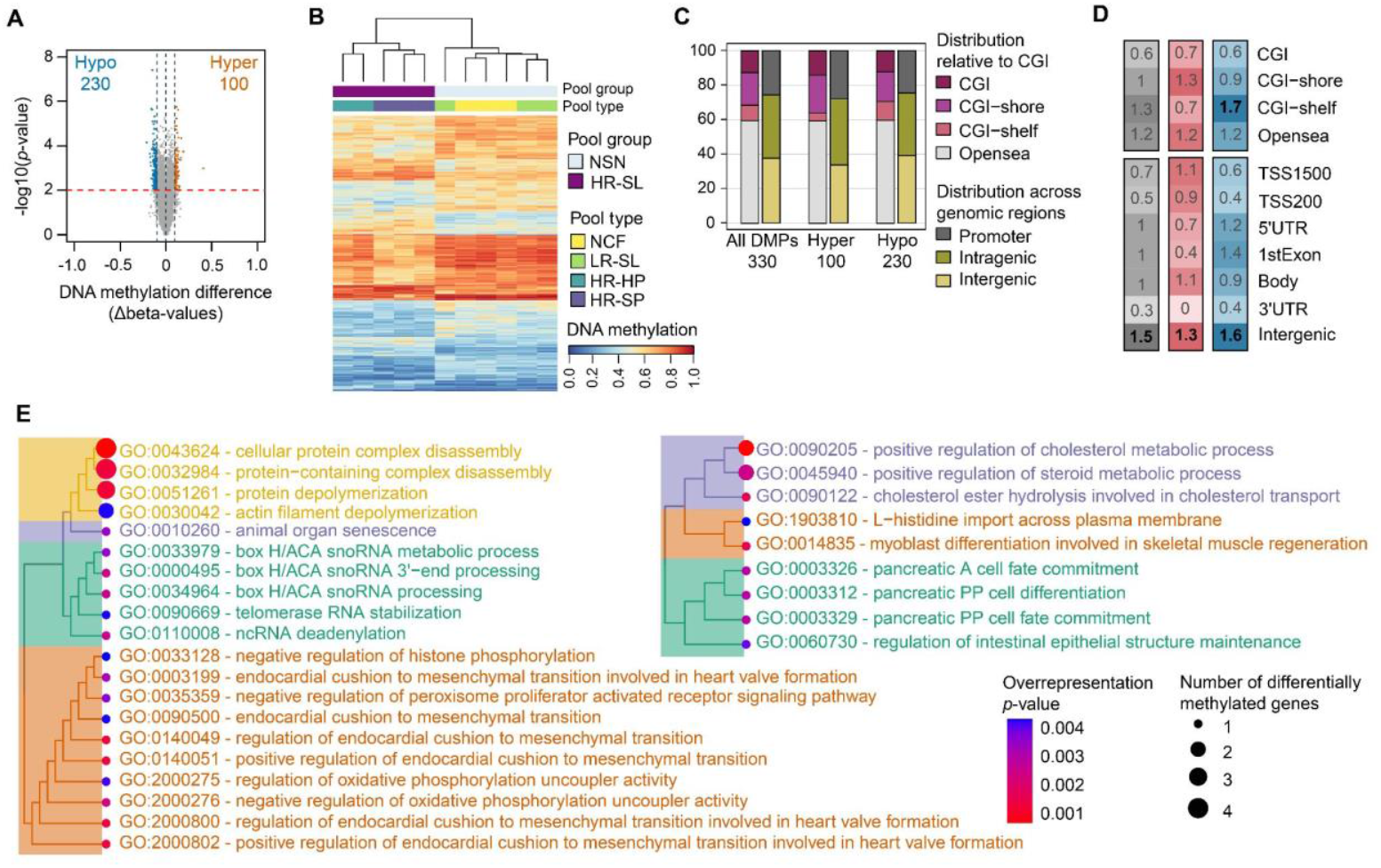
Characterization of the 330 differentially methylated positions (DMPs) between no serrated neoplasia and high-risk serrated lesions. **A**. Volcano plot showing the differential methylation -log_10_(*p*-value) for the 741,739 probes analyzed versus differences in methylation levels (Δbeta: obtained by subtracting the beta-values of NSN from HR-SL). Significant hypermethylated (Δbeta > 0.1) and hypomethylated (Δbeta < - 0.1) positions appear highlighted in orange or blue, respectively, above the red dashed line (*p*-value < 0.01). **B**. Hierarchical clustering and heatmap showing the methylation levels across the 13 cfDNA pooled samples for the 330 DMPs. Dendrograms were computed and reordered using Euclidean distance and a complete clustering agglomeration. Methylation levels (beta-values) range from 0 (blue, unmethylated) to 1 (red, fully methylated). **C**. Distribution of the 330 DMPs relative to CGI and functional genomic locations. **D**. Enrichment of 330 DMPs in relation to CGI annotation and functional genomic regions. The color scale indicates the fold enrichment of all DMPs (gray), hypermethylated (red), and hypomethylated (blue) positions. The bolded numbers indicate annotations that are enriched with respect to the distribution of probes on the MethylationEPIC array (odds-ratio > 1 and one-sided Fisher’s exact test *p*-value < 0.05). **E**. Hierarchical clustering of BP ontology terms based on semantic similarity. Treeplots of the 20 BP terms enriched in the hypermethylated promoter-associated DMPs (left) and 9 BP terms enriched in the hypomethylated promoter-associated DMPs. Nodes are colored with respect to the overrepresentation *p*-value and sized relatively to the number of differentially methylated genes annotated to each term. CGI (CpG island): region of at least 200 bp with a CG content > 50% and an observed-to-expected CpG ratio ≥ 0.6; CGI-shore: sequences 2 kb flanking the CGI, CGI-shelf: sequences 2 kb flanking shore regions, opensea: sequences located outside these regions, promoter regions (5′UTR, TSS200, TSS1500, and first exons), intragenic regions (gene body and 3′UTR), and intergenic regions. TSS200, TSS1500: 200 and 200-1500 bp upstream of the transcription start site, respectively. HR-HP: high-risk hyperplastic polyp, HR-SL: high-risk serrated lesion, HR-SP: high-risk serrated polyp, LR-SL: low-risk serrated lesion, NCF: no colorectal findings, NSN: no serrated neoplasia.

To identify biological functions annotated to the DMPs, we computed gene ontology (GO) enrichment analysis on the promoter-associated hypermethylated and hypomethylated DMPs separately. We identified 25 enriched GO terms associated with the DMPs that were hypermethylated in HR-SL compared to NSN. Differentially methylated genes overlapping with those GO terms were *PARN, TWIST1, COMP, ADD2, MICAL3, MRPL28, NCKAP5, CPLX2, RPEL1, CHST10*, and *LRRC26*. The hypomethylated DMPs reported the enrichment of 12 GO terms, including the genes *NEUROD1, WNT10B, CES1, FGF1, SLC7A1, NCEH1, GHR*, and *MRI1* (Supplementary Table 3). Within the BP (biological process) ontology, 20 GO terms were enriched in the hypermethylated DMPs, which were related to protein complex depolymerization and disassembly, animal organ senescence, box H/ACA snoRNA processing, and regulation of endocardial cushion to mesenchymal transition. On the other hand, 9 BP terms were enriched in the hypomethylated DMPs, related to the positive regulation of cholesterol metabolic process, L-histidine import across the membrane, and pancreatic cell fate commitment. Hierarchical clustering of BP terms according to semantic similarity is shown in Figure 1E.

Differential methylation analyses were also performed at region level, to test whether there were regional-specific differences in cfDNA associated with the different lesions belonging to the serrated pathway. Significant differentially methylated regions (DMRs) were defined as regions with at least two adjacent CpG sites yielding methylation differences in the same direction, with a FWER < 10%, and *p*-value < 0.01. Altogether, the results from all the comparisons performed rendered a total of 9 DMRs (Figure 2), some of them resulting significant in more than one comparison (Table 2). Two of these hypomethylated DMRs were identified comparing NSN and HR-SL: DMR1 annotated to the promoter (TSS200) of the *PRRT3* gene, and DMR2 located on the gene body of *NINJ2*. The largest methylation differences were found in DMR2 and DMR9 for the comparisons NCF vs HR-HP and HR-HP vs HR-SP, respectively.

**Table 2.**
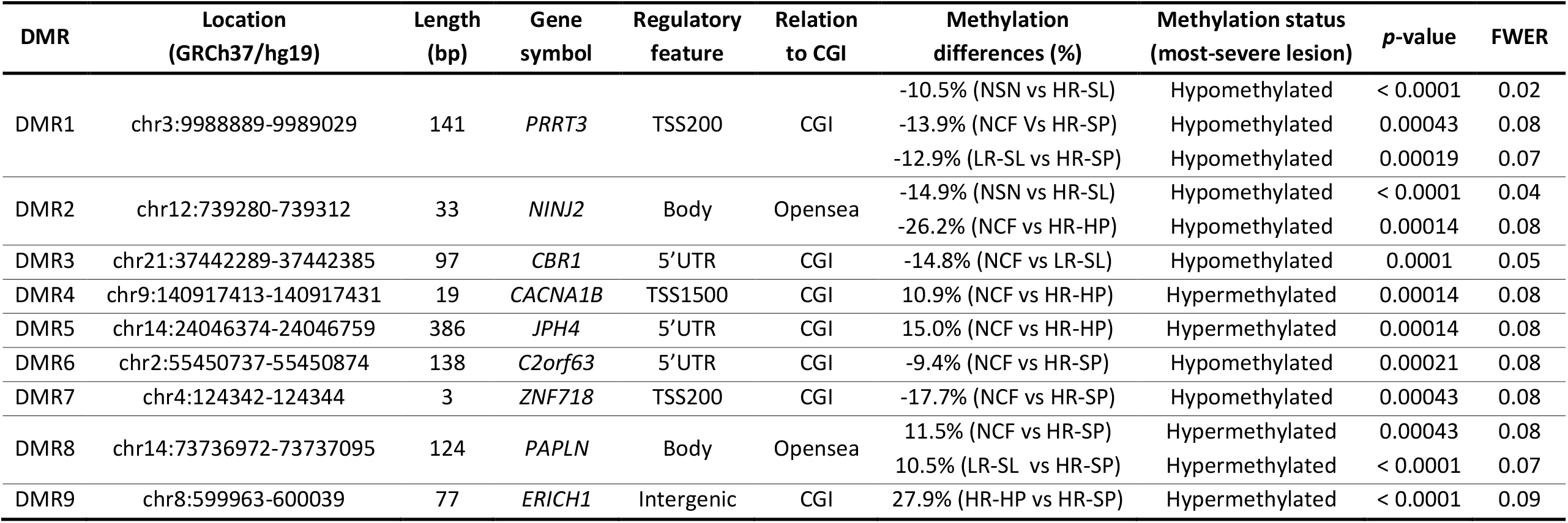
Differentially methylated regions. Annotation of the regions to regulatory features and CGI according to the Methylation EPIC Manifest. CGI: CpG island, region of at least 200 bp with a CG content > 50% and an observed-to-expected CpG ratio≥0.6; Opensea: sequences located outside CGI regions; Body: gene body (intragenic region); TSS200, TSS1500: 200 and 200-1500 bp upstream the transcription start site, respectively. FWER: family-wise error rate, HR-HP: high-risk hyperplastic polyp; HR-SL: high-risk serrated lesion; HR-SP: high-risk serrated polyp; LR-SL: low-risk serrated lesion; NCF: no colorectal findings; NSN: no serrated neoplasia.

**Figure 2.**
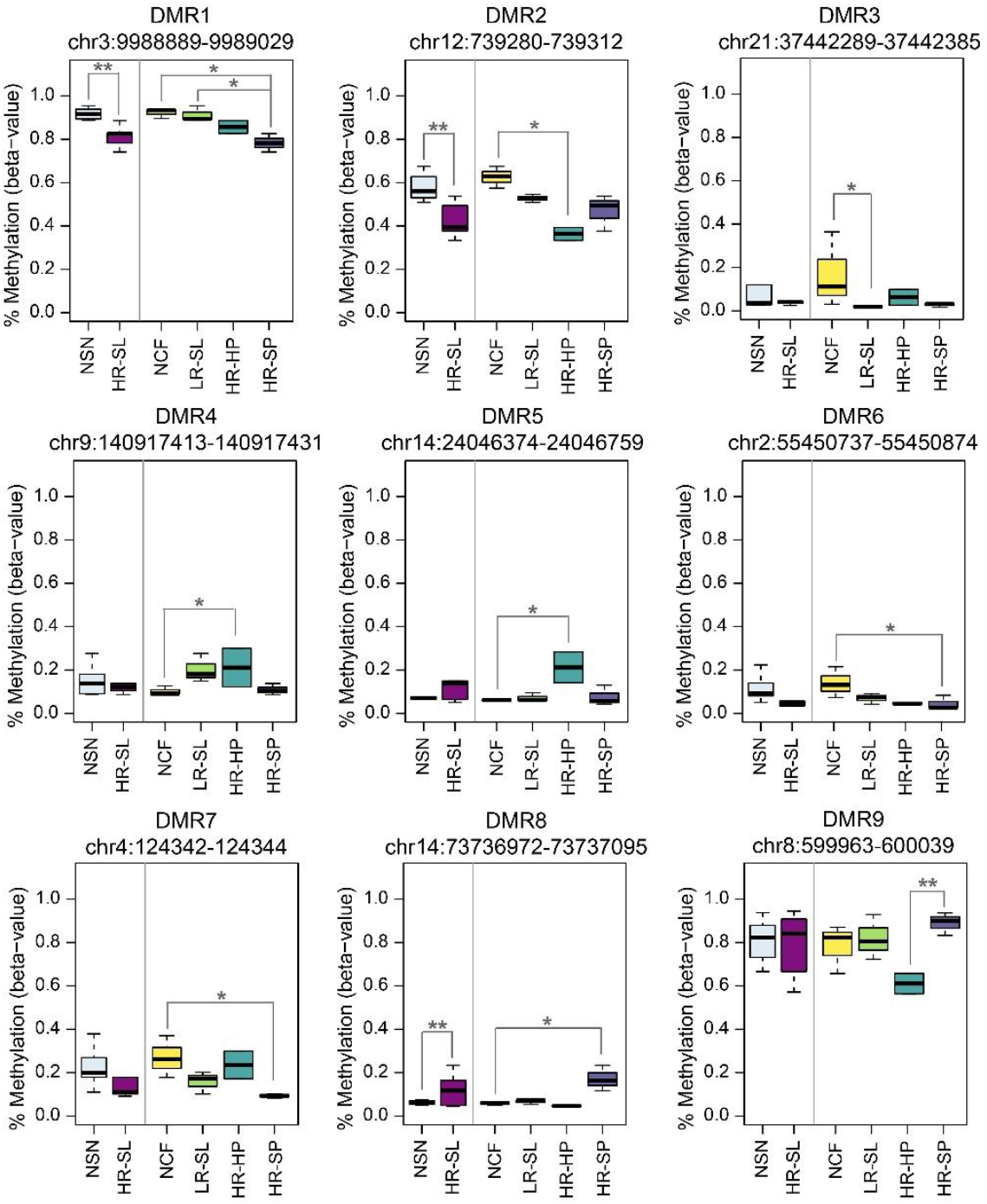
Methylation levels of the 9 differentially methylated regions (DMRs) in the cfDNA pooled samples. Methylation levels are shown as beta-values ranging from 0-unmethylated to 1-fully methylated (regional differential methylation \*\**p*-value < 0.0001; \**p*-value < 0.001). HR-HP: high-risk hyperplastic polyp, HR-SL: high-risk serrated lesion, HR-SP: high-risk serrated polyp, LR-SL: low-risk serrated lesion, NCF: no colorectal findings, NSN: no serrated neoplasia.

### Pathway-specific cfDNA methylome

The concordance between the differential methylation profile identified in serum cfDNA and tissue methylation was assessed. Two external datasets (GSE68060 and E-MTAB-7854)[22,23] were combined to obtain microarray methylation data from serrated CRC (n=38), SSL (n=13), and healthy mucosa (n=16) tissue samples. This exploration was restricted to 188 out of the 330 DMPs targeted by probes shared by the Methylation450k and MethylationEPIC arrays. The PCA and heatmap performed on SSL, serrated CRC and mucosa samples revealed that the differential methylation profile found between NSN and HR-SL cfDNA can clearly discriminate serrated pathological groups in tissue samples (Figure 3A and 3B).

**Figure 3.**
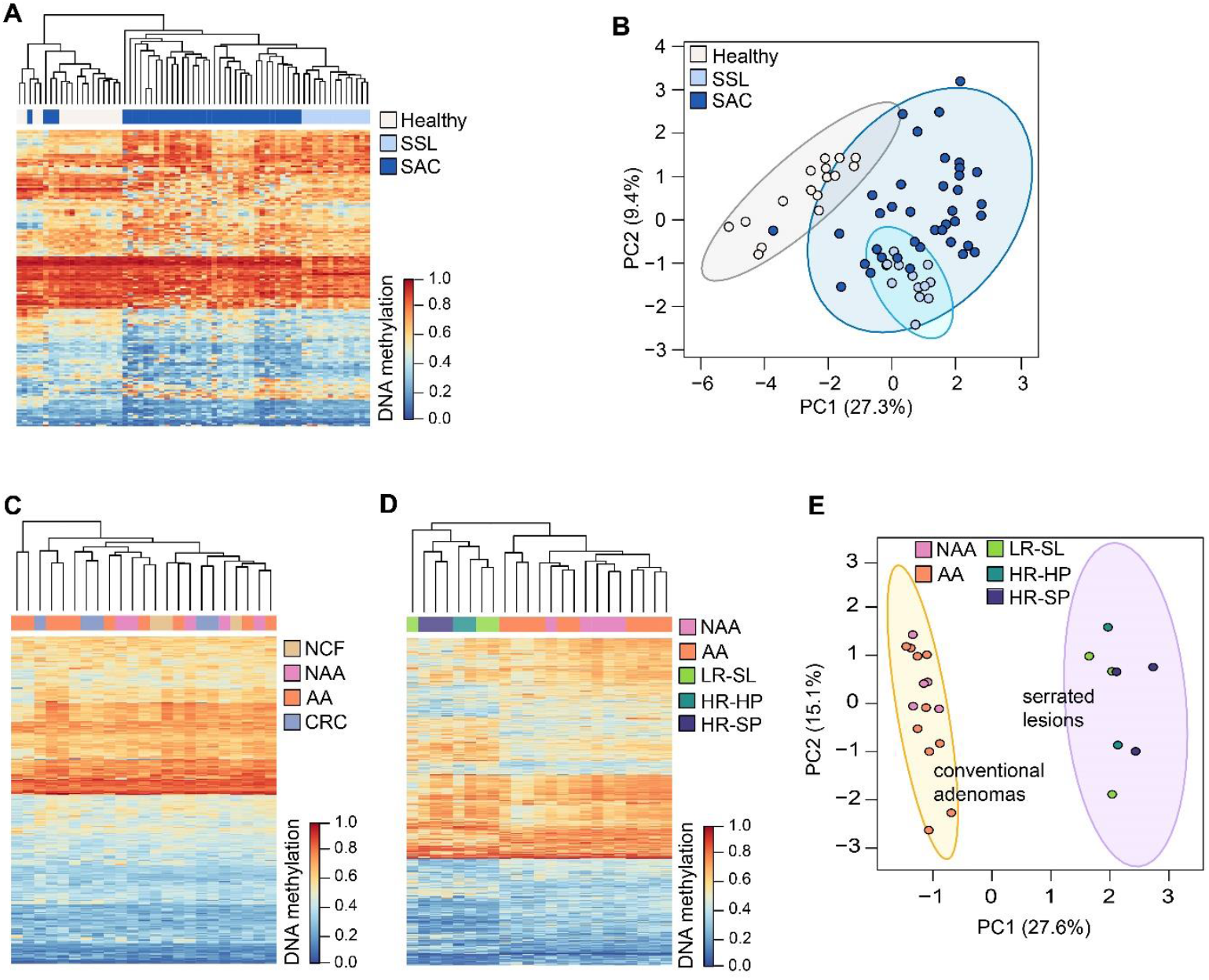
Differences in CRC pathways cfDNA methylation profiles. **A**. Heatmap and hierarchical clustering performed on tissue samples from the serrated pathway, based on the 188 DMPs shared by the 450k and EPIC arrays. **B**. PCA performed on tissue samples from the serrated pathway, based on the 188 DMPs shared by the 450k and EPIC arrays. **C**. Heatmap and hierarchical clustering showing the methylation levels of the 330 DMPs of the serrated pathway in 23 cfDNA pooled samples from the CRC conventional pathway. **D**. Heatmap and hierarchical clustering based on the 1,000 most-variable CpG positions in cfDNA pools. **E**. PCA based on the 1,000 most-variable CpG positions in cfDNA pools. For heatmaps, dendrograms were computed and reordered using Euclidean distance and a complete clustering agglomeration and methylation levels are expressed as beta-values ranging from 0 (blue, unmethylated) to 1 (red, fully methylated). PCA plots show the 95% confidence ellipses. AA: advanced adenomas, CRC: colorectal cancer, HR-HP: high-risk hyperplastic polyp, HR-SL: high-risk serrated lesion, HR-SP: high-risk serrated polyp, LR-SL: low-risk serrated lesion, NAA: non-advanced adenoma, NCF: no colorectal findings, SSL: sessile serrated lesions.

To check whether the cfDNA methylation profiles are specific of the different CRC carcinogenic pathways, cfDNA methylation data from the serrated pathway were compared with cfDNA methylation data from the conventional CRC pathway (GSE186381). This dataset includes MethylationEPIC data of cfDNA pooled samples of NCF (n=3), non-advanced adenomas (n=5), advanced adenomas (n=10), and CRC (n=5).[24] None of the 330 DMPs from the serrated pathway was differentially methylated in samples from the conventional pathway (10% FDR). Unsupervised clustering and heatmap from Figure 3C shows no difference in the methylation levels of the 330 DMPs from the serrated pathway and no ability to group advanced neoplasia samples from the conventional pathway. Just one CG, cg08779649 (chr13:50194554), met the criteria of *p*-value < 0.01 and at least 10% difference in the methylation levels. This CpG site is annotated to an opensea region located downstream of the CGI chr13:49092410-49092680. cg08779649 is hypermethylated (14.4%) in HR-SL, while hypomethylation (−13.8%) was observed in advanced neoplasia from the conventional pathway. Altogether these results suggest that the cfDNA differential methylation profile identified is specific to the serrated pathway. The pathway-specific differential methylation profile in serum can also be observed from early stages of carcinogenesis, as the 1,000 most-variables CpG sites display a different methylation profile between precursor lesions from both pathways (Figure 3D). The principal component analysis (PCA) based on the most-variable positions shows a clear separation between conventional adenomas (non-advanced and advanced) and serrated lesions (low-risk and high-risk) (Figure 3E).

### Evaluation of DMRs as potential biomarkers

MS-qPCR assays were successfully developed for DMR2, DMR7, and DMR9 (Supplementary Table 4). To explore the utility of the DMRs as biomarkers, their methylation status was evaluated in an independent cohort of 80 individual serum cfDNA samples (Table 1). The methylation levels of the DMRs analyzed did not follow a normal distribution. Methylation values of DMR7 were skewed towards 0%, while those of DMR2 and DMR9 were skewed towards 100%. DMR2, DMR7, and DMR9 were hypermethylated in HR-SL compared to NSN, with methylation differences of 2.4%, 5.8%, and 1.6%, respectively, although none of them was statistically significant. Statistically significant differences for DMR2 were found between NCF, LR-SL, and HR-HP vs SAC (methylation differences of 18.7%, 18.9%, and 37.9%, respectively) and between HR-HP and HR-SP (28.1%); between LR-SL and HR-HP for DMR7 (13.49%); and between HR-SP and SAC for DMR9 (−4.2%) (Figure 4A). No statistically significant differences were found for any of the DMRs between tumor tissue and matched healthy mucosa.

**Figure 4.**
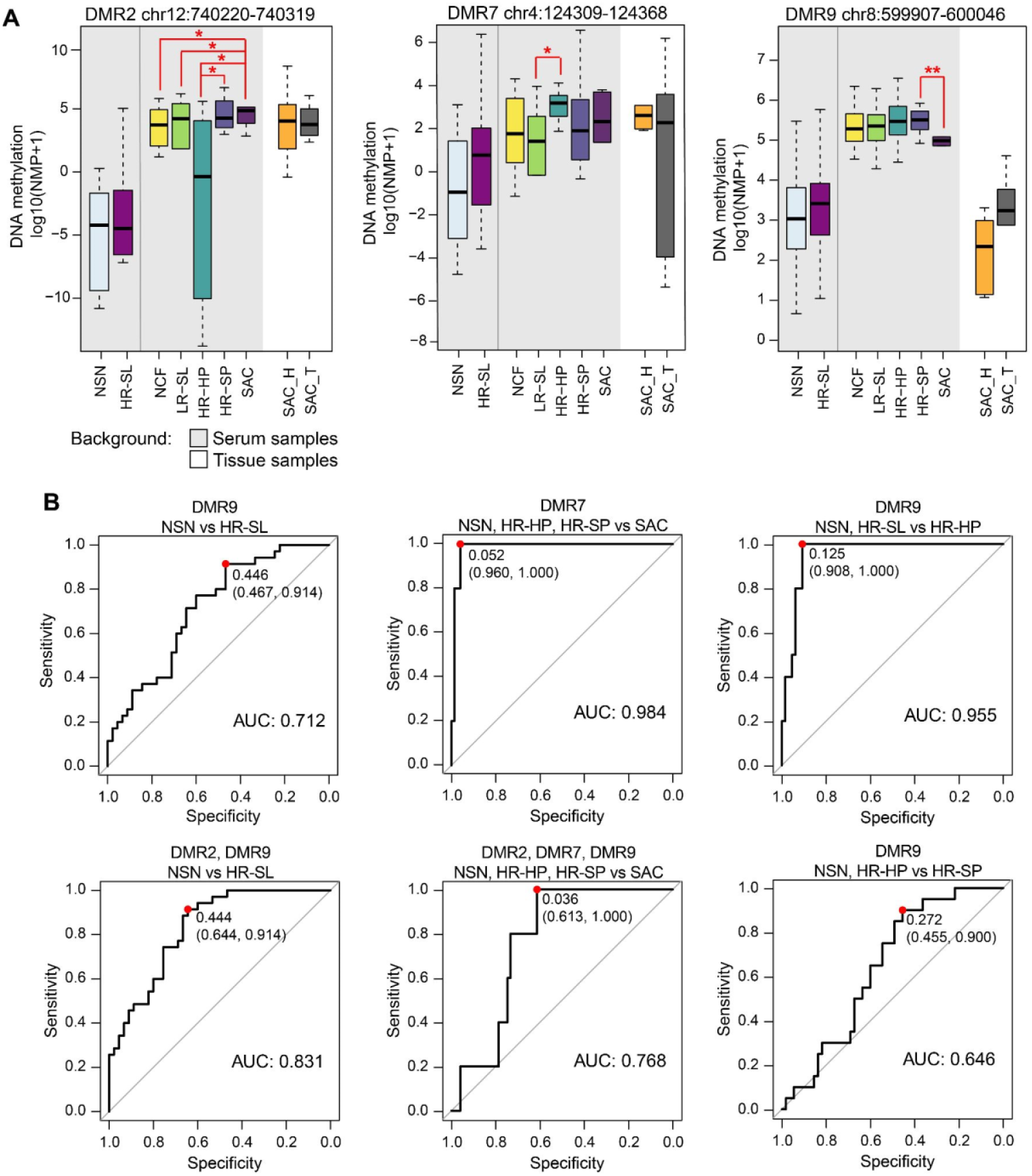
Targeted evaluation of the DMRs in serum. **A**. Methylation levels of the DMRs in individual serum cfDNA samples and matched SAC tumor-mucosa tissue samples, expressed as log10(NMP + 1) (*Wilcoxon rank-sum test *p*-value < 0.05, **Wilcoxon rank-sum test *p*-value < 0.01). **B**. ROC curve analysis and AUC for the best models obtained with single or combinations of DMRs for the detection of HR-SL, SAC, HR-HP, or HR-SP, derived by leave-one-out cross-validation in the individual serum samples (n=80). The red dots indicate the sensitivity and specificity values for the best cut-offs based on the Youden Index method. HR-HP: high-risk hyperplastic polyp, HR-SL: high-risk serrated lesion, HR-SP: high-risk serrated polyp, LR-SL: low-risk serrated lesion, NCF: no colorectal findings, NSN: no serrated neoplasia, SAC: serrated adenocarcinoma, SAC_H: healthy mucosa from SAC patients, SAC_T: tumor tissue from SAC patients.

Logistic regression models based on log10(NMP + 1) were elaborated for the detection of HR-SL (joint detection of HR-HP, HR-SP, and SAC) and HR-HP, HR-SP, and SAC separately. The discriminatory capacity of individual or combined DMRs was assessed by ROC curve analysis and leave-one-out cross-validation (Figure 4B and Supplementary Figure 4). Results from the best performing models are summarized in Table 3. The combination of DMR2 and DMR9 discriminated HR-SL from NSN with 64.4% specificity and 91.4% sensitivity (AUC 0.831, 95% CI: 0.745-0.917), and detected all the SAC cases. DMR7 also discriminated all the patients with SAC (100% sensitivity) from patients with NSN, HR-HP and HR-SL (96% specificity) (AUC 0.984, 95% CI: 0.959-1). Regarding models for the discrimination of precancerous lesions, DMR9 showed the best performance for the detection of HR-HP and HR-SP, yielding 90.7% specificity and 100% sensitivity (AUC 0.955) for HR-HP, while HR-SP were identified with 45.5% specificity and 90% sensitivity (AUC 0.646).

**Table 3.** Performance of the DMRs as biomarkers for serrated lesions in 80 individual serum samples with the corresponding 95% confidence intervals (95% CI). The detection rates of HR-SL and SAC are also shown. No significant differences were found between the detection of distal versus proximal lesions for all models (Fisher’s exact test p-value>0.05). ROC curves and performance parameters were derived by the leave-one-out cross-validation approach. AUC: area under the curve, HR-HP: high-risk hyperplastic polyp, HR-SL: high-risk serrated lesion, HR-SP: high-risk serrated polyp, NSN: no serrated neoplasia, SAC: serrated adenocarcinoma

|  |  | Model performance |  |  | Detection rates (%) |  |  |  |  |  |  |
| --- | --- | --- | --- | --- | --- | --- | --- | --- | --- | --- | --- |
|  |  | AUC<br>(95% CI) | Specificity %<br>(95% CI) | Sensitivity %<br>(95% CI) | HR-HP<br>distal | HR-HP<br>proximal | All | HR-SP<br>distal | HR-SP<br>proximal | All | SAC |
| NSN vs HR-SL | DMR9 | 0.712<br>(0.601–0.825) | 46.7<br>(32–62) | 91.43<br>(77–98) | 100 | 60 | 80 | 100 | 91.7 | 95 | 100 |
|  | DMR2, DMR9 | 0.831<br>(0.745–0.917) | 64.4<br>(49–78) | 91.4<br>(77–98) | 100 | 80 | 90 | 100 | 83.3 | 90 | 100 |
| NSN, HR-HP, HR-SL vs SAC | DMR7 | 0.984<br>(0.959–1) | 96<br>(89–99) | 100<br>(48–100) | - | - | - | - | - | - | 100 |
|  | DMR2, DMR7, DMR9 | 0.768<br>(0.633–0.903) | 61.3<br>(49–72) | 100<br>(48–100) | - | - | - | - | - | - | 100 |
| NSN, HR-SP vs HR-HP | DMR9 | 0.955<br>(0.912–0.999) | 90.7<br>(81–97) | 100<br>(69–100) | 100 | 100 | 100 | - | - | - | - |
| NSN, HR-HP vs HR-SP | DMR9 | 0.646<br>(0.520–0.773) | 45.5<br>(32–59) | 90<br>(68–99) | - | - | - | 100 | 83.3 | 90 | - |

## DISCUSSION

Although CRC represents the classical model of epithelial neoplasm development through the so-called adenoma-to-carcinoma sequence,[25] it is now well established that there are many molecular mechanisms underlying this sequence that drive CRC development.[26] The serrated neoplasia pathway accounts for 15-30% of CRC cases,[2–4] with serrated adenocarcinomas having a worse prognosis compared to conventional CRC due to weak immune response, high tumor budding and an infiltrative tumor pattern.[27,28]

The early detection of CRC and precancerous lesions is determinant for the success of screening programs. Several biological characteristics of the serrated lesions have clinical relevance for the screening and colonoscopic surveillance of SAC. Thus, with the serrated pathway becoming widely recognized,[6,29] an accurate early detection of serrated lesions could reduce both the incidence of SAC and the serrated-lesion related interval CRC. To the best of our knowledge, this study explored for the first time the serum methylome in patients with precursor lesions from the serrated pathway, aiming its characterization and evaluation of cfDNA methylation as a potential source of biomarkers for the non-invasive screening and diagnosis of the serrated pathway.

First, we conducted an epigenome-wide methylation analysis of serum cfDNA, combining the MethylationEPIC array with a sample pooling approach. We identified a cfDNA differential methylation profile between HR-SL (large HP, large SSL, SSL with dysplasia, and TSA) and NSN (individuals with no colorectal findings, small HP, and small non-dysplastic SSL and SP), reporting 330 DMPs of which 39.1% are associated to CpG islands. All the different serrated lesions included were pairwise compared to obtain 9 significant DMRs, of which 7 were located within the CpG islands of the genes *PRRT3, CBR1, CACNA1B, JPH4, C2orf63, ZNF718*, and *ERICH1*.

Several high-throughput methylation analyses of tissue samples have identified the gradual accumulation of methylation changes at all steps of the progression of the serrated pathway,[10,13,23,30] reporting significant hypomethylation even at higher frequency than hypermethylation. This is consistent with the differential methylation profile we report in serum cfDNA, where 69.7% of the DMPs were hypomethylated in HR-SL and DMRs were mostly hypomethylated in the most-severe lesion. The serum cfDNA methylation profile was explored in tissue samples from healthy mucosa, SSL and SAC, combining external methylation microarray data.[22,23] The differential methylation pattern found between HR-SL and NSN cfDNA can also discriminate serrated pathological groups in tissue samples. Though this verification is limited, some degree of concordance between serum and tissue methylation can be observed. It is worth mentioning that discrepancy in the frequencies of methylation alterations in tumor tissue and cfDNA has been reported, showing the latter considerably lower frequencies.[31]

Different oncogene mutation and expression profiles, MSI status, methylome signatures, and epigenetic regulation have been observed between SAC and conventional and sporadic CRC tissue samples.[22,32,33] Here we evidenced that the serum differential methylation profile is also pathway-specific, as the DMPs identified for HR-SL have no ability to distinguish conventional colorectal neoplasia in cfDNA pooled samples. Moreover, when pooled cfDNA samples from both pathways are merged, the most-variable CpG sites exhibit different methylation levels in conventional compared to serrated precursor lesions, suggesting that pathway-specific serum methylation profiles can already be detected from early stages of carcinogenesis.

We also made a preliminary evaluation of three DMRs as non-invasive biomarkers for the detection of HR-SL and SAC in an independent cohort of individual serum samples. The methylation levels were quantified in serum and tissue samples, targeting DMR2 (chr12:740220-740319) located in the body of *NINJ2*, DMR7 (chr4:124309-124368) located on the CpG island of *ZNF718*, and DMR9 (chr8:599907-600046) located on the CpG island of *ERICH1*. Logistic regression models based on unique or combinations of DMRs were cross-validated to derive classification rules for the detection of serrated lesions in serum. The combination of DMR2 and DMR9 detected HR-SL (HR-HP, HR-SP, and SAC) with 91.4% sensitivity and 64.4% specificity, and reported detection rates of 90%, 90%, and 100%, for HR-HP, HR-SP, and SAC, respectively. For the discrimination of SAC from all the other lesions, DMR7 showed 100% sensitivity and 96% specificity. We also explored the ability of the DMRs to specifically detect precursor lesions. DMR9 showed the best performance for the independent detection of HR-HP and HR-SL, reporting sensitivities of 100% and 90% for HR-HP and HR-SP, respectively, at 90.7% and 45.5% specificity. No statistically significant differences were found for any of the DMRs between tumor tissue and matched healthy mucosa, probably due to small sample size (n=6).

The morphology of serrated lesions makes them less prone to bleed, which limits the sensitivity of FIT for detecting SSL regardless of their size and grade of dysplasia.[17–19,34] The sensitivities showed by our DMRs are superior to that of FIT for the detection of serrated lesions, which ranges from 6.2-20.4% for SSL at 87.4-96.8% specificity,[17,18] and shows 7.4% sensitivity for HR-SL when the specificity is fixed to 95%.[19] Although most screening programs are based on FIT, the aforementioned studies suggest its limited value to detect serrated lesions, resulting ineffective to prevent interval cancers arising from these lesions.

A few studies have evaluated the performance of DNA methylation non-invasive tests for the detection of precursory serrated lesions. The only blood test approved by the FDA for the detection of CRC is based on *SEPT9* methylation.[35] In an opportunistic screening study, the plasma *SEPT9* assay showed a sensitivity of 27.8% for the detection of serrated polyps, 21.3% for HP, and 40.9% for conventional adenomas, at 78.4% specificity.[36] Another study evaluating methylation of *BCAT1*/*IKZF1* in plasma reported a sensitivity of 8.8% for the detection of SSL at 93% specificity.[18] The FDA-approved multitarget fecal test detected serrated lesions with 40.7-42.4% sensitivity and 86.6-89.1% specificity.[19,34] The cross-validated diagnostic performance of the DMRs evaluated outperformed other methylation non-invasive tests for the detection of serrated lesions, adding more evidence to the previous observations that DNA methylation is more sensitive for serrated lesions than FIT. This fact should be discussed when it comes to the non-invasive screening of serrated CRC.

It is worth mentioning that our multicentre cohort includes the whole pathological range of the serrated pathway, from small serrated and hyperplastic polyps, to TSA and large and dysplastic SSL, as well as SAC and colonoscopically confirmed healthy controls. The sample size is justified by the low prevalence of serrated lesions in colonoscopy screening, with colonoscopy detection rates of 1.8% for large HP, 0.8-4.6% for SSL, 0.8-1.6% for large or dysplastic SSL, and 0.2-4.4% for TSA.[37,38] In relation to CRC, its prevalence in colonoscopic studies ranges from 0.28-0.42% for all types of CRC.[39,40] As the serrated pathway accounts for up to 30% of CRC, the small number of SAC serum samples available for our study was expected. This precluded the construction of cfDNA pooled samples from SAC cases, that therefore were retained for the biomarker evaluation phase.

As far as we are concerned, this is the only study available to date exploring the serum methylome of precursory lesions of the serrated pathway. We have reported a differential methylation profile that can distinguish HR-SL from NSN, showing concordance with tissue methylation from different external datasets. The methylation profiles in serum cfDNA are pathway-specific, and may serve as a source of non-invasive biomarkers for the detection of HR-SL and SAC in screening programs.

## METHODS

### Patient characteristics and samples

Individuals were recruited from the following Spanish Hospitals: Hospital Donostia (San Sebastián), Hospital Clínic de Barcelona (Barcelona), Hospital General Universitario de Alicante (Alicante), Complexo Hospitalario Universitario de Ourense (Ourense), Hospital Álvaro Cunqueiro (Vigo). Individuals with incomplete colonoscopy or suboptimal bowel preparation, personal history of CRC, digestive cancer, inflammatory bowel disease, serrated polyposis syndrome, previous colectomy, or with a severe synchronic illness were excluded. The age range of the patients matches the USPSTF guideline recommendation for CRC screening (50-75 years).[41,42]

All individuals underwent a colonoscopy, performed by experienced endoscopists following the recommendations from the Spanish guidelines on quality of colonoscopy.[43] Blood samples were obtained before the colonoscopy procedure from 186 individuals between 51-76 years old. Serum was collected after coagulation and centrifugation and was stored at −20 °C until cfDNA extraction.

Individuals were classified according to the most advanced colorectal finding.[44] We grouped samples into five main categories: (i) SAC (including CRC with MSI, CIMP-high, or BRAF mutation; mucinous adenocarcinoma and signet-ring cell adenocarcinoma), (ii) HR-SP (comprising TSA, SSL, and SP with dysplasia or ≥ 10 mm), (iii) HR-HP (HP ≥ 10 mm), (iv) LR-SL (SP without dysplasia < 10mm and HP < 10mm), and (v) NCF. Serum samples were separated into two independent subsets: 106 serum samples (30 NCF, 30 LR-SL, 16 HR-HP, and 30 HR-SP) for the genome-wide methylation analysis and 80 samples (20 NCF, 25 LR-SL, 10 HR-HP, 20 HR-SP, and 5 SAC) for the targeted evaluation of methylation biomarkers. Formalin-fixed paraffin-embedded (FFPE) tumor tissue from 6 SAC cases and matched normal mucosa from the same patient were also used to evaluate the methylation biomarkers (Table 1).

All individuals provided written informed consent, and the study followed the ethical and clinical practices of the Spanish Government and the Helsinki Declaration, and was approved by the Galician Ethical Committee for Clinical Research (2018/008).

### cfDNA isolation, sample pooling and DNA extraction from FFPE samples

cfDNA was extracted from 0.5-2 mL serum, according to availability. For the epigenome-wide analysis we followed a sample pooling approach. First, cfDNA was isolated from serum samples with a phenol-chloroform protocol[45] and was quantified with the Qubit dsDNA HS Assay Kit (Thermo Fisher Scientific, Waltham, MA, USA). Then, 11 independent pooled samples were prepared combining equal amounts of cfDNA from 10 individuals (half male, half female) as previously described.[20] The factors considered to match between pooled samples were sex, age, and recruitment hospital (Supplementary Table 1). Pooled samples were stored at -20°C.

For the targeted evaluation, cfDNA was extracted from serum samples with the QIAmp Circulating Nucleic Acids Kit (Qiagen, Hilden, Germany). DNA was extracted from FFPE tissue specimens with the NucleoSpin® DNA FFPE XS DNA Isolation kit (Macherey-Nagel, Allentown, PA, USA) with xylene deparaffinization. DNA and cfDNA samples were bisulfite treated with the EZ DNA Methylation-Direct Kit (Zymo Research, Irvine, CA, USA) and stored at -80°C.

### Genome-wide methylation profiling of cfDNA

The cfDNA pooled samples (n=11) were stored at − 20 °C and sent to the Josep Carreras Leukaemia Research Institute (Badalona, Spain) for processing and methylation quantification. Pools were bisulfite-treated in the same batch and were hybridized to Infinium MethylationEPIC BeadChip arrays (Illumina, San Diego, CA, USA) following the manufacturer’s instructions. Samples from different pathological groups were carefully allocated to each slide to minimize confounder variability due to technical batch effects. Methylation levels were derived from a total of 866,091 CpG sites.

Illumina methylation data were preprocessed and analyzed using the R environment (versions 3.3.3 and 3.4.0) with R and Bioconductor packages. Microarray dye-bias and technical batch effects were corrected by the single-sample out-of-band normalization and the *ComBat* method. Poor quality and cross-reactive probes, probes annotated to genetic variants and probes targeting X/Y chromosomes were discarded. A total of 734,739 CpG sites —mapped to the human genome assembly GRCh37/hg19 and annotated to CGI, promoter, intragenic and intergenic regions— remained for analysis. No samples were removed due to quality issues (see Supplementary Methods for details).

### Differential methylation analyses

Differentially methylated CpG positions (DMPs) were identified by the standard workflow of the *limma* package:[46] linear models were fitted for each CpG and *p*-values were computed with an empirical Bayes moderated t-test. A *p*-value < 0.01 and at least 10% difference in the methylation levels were used as the threshold to select DMPs. We identified differentially methylated regions (DMRs) by linear regression and sample permutation applying the *bumphunter* method.[47] The false positive rate was controlled by the family-wise error rate (FWER). Significant DMRs were selected as those with FWER < 10%, *p*-value < 0.01, and with at least two adjacent CpG sites.

Serrated pathway differential methylation profiles were explored in cfDNA pooled samples from the conventional CRC pathway (GSE186381). This dataset includes 23 cfDNA pooled samples grouped in NCF, non-advanced adenomas, advanced adenomas and CRC.[24] Serum differential methylation profiles were also tested in tissue samples from healthy colorectal mucosa (n=16) and serrated tumors (n=38) from the dataset GSE68060,[22] and from SSL (n=13) from the dataset E-MTAB-7854.[23] This analysis was limited to probes shared by the Methylation450k and MethyationEPIC BeadChip arrays.

### Functional annotation of differential methylation

One-sided Fisher’s exact tests were used to assess the significance of the enrichment of the DMPs to functionally annotated elements (CGI, promoter, intergenic and intragenic regions). To determine the biological functions of the DMPs associated with serrated neoplasia, gene ontologies (GO) enrichment analysis were conducted using *gometh*.[48] Significantly enriched GO terms were obtained for promoter regions based on hypomethylated and hypermethylated DMPs separately. A *p*-value < 0.005 was considered statistically significant. GO term semantic similarity analyses were performed based on the Jaccard’s similarity index.[49]

### Targeted evaluation of differentially methylated regions

Nested custom qPCR assays were used for targeted methylation analyses in serum (n=81) and tissue samples (n=12). First, bisulfite-treated cfDNA or DNA was subjected to a pre-amplification (pre-PCR) with primers flanking the region of interest, followed by a MS-qPCR (methylation-specific qPCR) with a probe targeting the methylated sequence, using diluted pre-amplification products as template (see Supplementary Table 4 and Supplementary Methods for details on primers and PCR conditions). A fully methylated control and a fully unmethylated control were included in each run for normalization and to verify plate-to-plate consistency. Standard curves were elaborated for each amplicon of interest using dilutions of the fully methylated control (100-0.01% methylation; amplification efficiency > 90%, R^2^ > 0.99). Raw methylation percentages (RMP) were estimated based on the corresponding standard curve. Then, RMP were normalized for DNA input, obtained for each sample by targeting a methylation independent region of the β-actin gene (*ACTB*). Normalized methylation percentages (NMP) were calculated as follows:

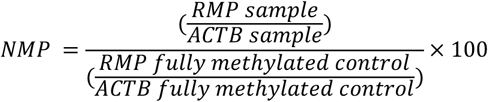

For data analysis, methylation values were transformed to log10(NMP + 1). Methylation levels were compared with the Wilcoxon rank-sum test for individual serum samples, and the non-parametric Wilcoxon signed-rank test for matched tumor and healthy mucosa tissue samples. Multivariate logistic regressions were fitted to the NMP to derive classification models for the detection of HR-HP, HR-SP, and SAC. The classification performance was assessed by leave-one-out cross validation, receiver-operating characteristic (ROC) curves were elaborated, and AUC, sensitivity, and specificity values were estimated with their corresponding 95% confidence intervals. The Youden Index method was used to determine the best cut-off values.[50] Fisher’s exact tests were employed to compare the proportion of distal and proximal lesions detected. All statistical analyses were performed with the R environment (version 3.4.0).

## Data Availability

The Infinium MethylationEPIC data from all the pooled samples generated and analyzed during this study have been deposited in the NCBI Gene Expression Omnibus (GEO) (www.ncbi.nlm.nih.gov/geo) and are accessible through GEO Series accession number GSE199173.

https://www.ncbi.nlm.nih.gov/geo/query/acc.cgi?acc=GSE199173

## ABBREVIATIONS

AA: advanced adenoma
AUC: area under the curve
cfDNA: circulating cell-free DNA
CRC: colorectal cancer
CGI: CpG island
DMR: differentially methylated region
DMP: differentially methylated position
FDR: false discovery rate
FIT: fecal immunochemical test
FWER: family-wise error rate
HP: hyperplastic polyp
HR-HP: high-risk hyperplastic polyp
HR-SL: high-risk serrated lesion
HR-SP: high-risk serrated polyp
MS-qPCR: methylation-specific quantitative PCR
NAA: non-advanced adenomas
NCF: no colorectal findings
NMP: normalized methylation percentage
NSN: no serrated neoplasia
RMP: raw methylation percentage
ROC: Receiver-operating characteristic
SAC: serrated adenocarcinoma
SSL: sessile serrated lesion
SP: serrated polyp
TSA: traditional serrated adenoma.

## DECLARATIONS

### Ethics approval and consent to participate

Written informed consent was obtained from all patients with approval by the Ethics Committee for Clinical Research of Galicia (2018/008). The study was conducted according to the clinical and ethical principles of the Spanish Government and the Declaration of Helsinki.

### Author’ contributions

LD conceived and designed the study. LD, JC, and ME supervised the study. MGG, LCR, LD, LAR, and CAGP contributed to the sample preparation and data acquisition. MGG and LD performed the analysis and interpretation of data. MGG, LCR and LD contributed to the experimental design. JC, LB, MB, AC, FB, RJ, ATB, and JGCF provided clinical advice for the study design, collection, and management of clinical data. MGG and LD prepared the manuscript. All authors critically reviewed and approved the final manuscript.

## Acknowledgments

We would like to thank Dr. Martínez Zorzano, and Professors Rodríguez Berrocal and Páez de la Cadena for their support and scientific advice. The samples and clinical data of patients included in this study were provided by: the HGUA Biobank (PT13 / 0010/0044), integrated into the *Red Nacional de Biobancos* and in the *Red Valenciana de Biobancos;* Basque Biobank/Biodonostia Node; Biobank at the Galicia Sur Health Research Institute; and Biobank of Hospital Clínic, Barcelona - IDIBAPS, with the approval of the respective Ethical and Scientific Committees, and have been processed following standard procedures. We acknowledge CESGA (Fundación Pública Galega Centro Tecnolóxico de Supercomputación de Galicia) for providing access to computing facilities to analyze methylation microarray data. Figure color palettes were adapted from Blake Robert Mills (2022). MetBrewer: Color Palettes Inspired by Works at the Metropolitan Museum of Art. R package version 0.1.0.

