## Supplementary for "Serum DNA methylome of the colorectal cancer serrated pathway"

**SUPPLEMENTARY METHODS**

**Bioinformatics preprocessing of microarray methylation data**

Illumina methylation data were preprocessed and analyzed using the R environment (versions 3.3.3 and 3.4.0) with R and Bioconductor packages. Raw methylated and unmethylated signal intensities were background and dye-bias corrected applying a normal-exponential convolution based on the non-specific fluorescence of Infinium I probes (single-sample out-of-band normalization method implemented in the *minfi* package).[1] The technical BeadChip batch effect was adjusted by an empirical Bayes method implemented in the *ComBat* function from the *sva* package.[2] The DNA methylation level was summarized for each CpG probe as the fraction of the methylated signal intensity over the total signal intensity (beta-value). Detection *p*-values were computed with the *minfi* package. Probes with a *p*-value > 0.01 and a bead count < 3 were discarded, and mean detection *p*-values were examined across all samples to identify any failed sample. After excluding cross-reactive probes,[3] probes violating any assumption for linear regression model fitting (linearity, homoscedasticity, uncorrelatedness, and normality of the standardized residuals),[4] probes annotated with genetic variants, and those located on the X and Y chromosomes, a total of 734,739 CpG sites remained for analysis. No samples were removed due to quality issues.

Probes were mapped to the human genome assembly GRCh37/hg19, annotated to genomic regions, and RefSeq genes according to the MethylationEPIC Manifest. CpG sites were assigned to CpG island (CGI), CGI-shores and shelves or opensea regions. CpG sites were also annotated as in promoter regions (5'UTR, TSS200, TSS1500, and first exons), intragenic regions (gene body and 3'UTR), and intergenic regions. Beta-values (methylation levels) were reported for interpretability and *M*-values (logit-transformation of beta-values) were used for statistical analyses, as they approximate a Gaussian distribution.[5]

**Differential methylation analyses**

Differentially methylated CpG positions (DMPs) were identified by the standard workflow of the *limma* package.[6] Briefly, linear models were fitted for each CpG across all samples by generalized least squares, and an empirical Bayes moderated t-test was used to compute the *p*-values. A *p*-value < 0.01 and at least 10% difference in the methylation levels were used as the threshold to select DMPs. We identified differentially methylated regions (DMRs) using the *bumphunter* method,[7,8] which searches for contiguous CpG sites consistently hypomethylated or hypermethylated between groups. Clusters were defined by neighboring probes within a window of 250 bp, then linear regressions were fitted to each probe, and 100 permutations were used to generate a null distribution of regions for establishing significance. The false positive rate was controlled by the family-wise error rate (FWER), a more conservative approach compared with the BH and FDR methods.[9] Significant DMRs were selected as those with FWER < 10%, *p*-value < 0.01, and with at least two adjacent CpG sites.

**Functional annotation of differential methylation**

One-sided Fisher's exact tests were used to assess the significance of the enrichment of the DMPs to functionally annotated elements such as CpG islands, promoter regions, and intergenic regions, using the annotation of the complete array as background. To determine the biological functions of the DMPs associated with serrated neoplasia, we conducted gene ontologies (GO) enrichment analyses using the *gometh* function from the *missMethyl* package,[10] which accounts for the bias derived from the differing number of probes targeting each gene and from probes annotated to multiple genes, applying a Wallenius' non-central hypergeometric test. Significantly enriched GO terms were obtained for promoter regions based on hypomethylated and hypermethylated DMPs separately. Overrepresentation results within a $p$-value < 0.005 were considered statistically significant. Biological process (BP), cellular component (CC), and molecular function (MF) ontologies were included. GO term semantic similarity analyses were performed with the *enrichplot* package[11] using the Jaccard's similarity index.[12]

**MS-qPCR analysis and fully methylated/unmethylated controls**

Pre-PCR reactions were performed in a 25 µL reaction mix containing 2 µL of bisulfite-modified DNA, 0.72 µM forward and reverse primers, a 75 µM dNTPs mixture, a 1X Ex Taq Buffer, and 1 unit of Takara Ex Taq HotStart (Takara Bio Inc., Kusatsu, Japan), with the following cycling conditions: 95 °C for 5 min, 25 cycles of 95 °C for 30 s, 30 s at 60 °C for all amplicons, 72 °C for 30 s, and finally 72 °C for 7 min. MS-qPCR was performed using the following dilutions of the previous PCR product: 1/20 for DMR2, DMR9 and ACTB, and 1/200 for DMR7. Real-time PCR was carried out in triplicate in a 20 µL volume containing 2 µL of the diluted pre-PCR, 600 nM of each primer, 200 nM of probe, and 1X TaqMan Gene Expression Master Mix (Thermo Fisher Scientific, Waltham, MA, USA), with an annealing temperature of 60 °C for DMR7, DMR9 and ACTB, and of 58 °C for DMR2, during 40 cycles. Amplifications were carried out in 96-well plates and run on a StepOne Plus instrument (Thermo Fisher Scientific, Waltham, MA, USA).

A fully methylated control (methyltransferase-treated DNA with M.SssI; New England Biolabs, Ipswich, MA, USA) and a fully unmethylated control (whole-genome amplification of DNA with GenomiPhi V2 DNA Amplification kit; GE Healthcare, Chicago, IL, USA) were included in each run for normalization and to verify plate-to-plate consistency. DNA extracted from peripheral blood from a donor was used to prepare controls.

**SUPPLEMENTARY TABLES**

**Supplementary table 1. Description of cfDNA pooled samples**.

| Pool type | Age median (range) | Total amount DNA (ng) | Pathology description |
|---|---|---|---|
| NCF | 61 (53-73) | 247.8 | Each pool contained 10 individuals with NCF |
| | 61.5 (53-71) | 172.8 | |
| | 63 (52-72) | 123.9 | |
| LR-SL | 61.5 (54-74) | 269.9 | 9 Individuals with HP < 10 mm, 1 individual with a SSP < 10 mm |
| | 61 (51-71) | 328.0 | 9 Individuals with HP < 10 mm, 1 individual with a SSP < 10 mm |
| | 61.5 (52-71) | 225.6 | 9 Individuals with HP < 10 mm, 1 individual with a SSP < 10 mm |
| HR-HP | 62.5 (54-68) | 259.8 | 8 Individuals with HP > 10 mm |
| | 61.5 (52-71) | 330.2 | 8 Individuals with HP > 10 mm |
| HR-SP | 61.5 (54-70) | 174.4 | 7 Individuals with dysplastic SSP, 2 individuals with SSP > 10 mm, 1 individual with TSA |
| | 63 (51-71) | 298.2 | 6 Individuals with dysplastic SSP, 2 individuals with SSP > 10 mm, 2 individual with TSA |
| | 62.5 (53-71) | 336.0 | 4 Individuals with dysplastic SSP, 5 individuals with SSP > 10 mm, 1 individual with TSA |

NCF: no colorectal findings; HP: hyperplastic polyps, HR-HP: high-risk hyperplastic polyp, HR-SP: high-risk serrated polyp, LR-SL: low-risk serrated lesion, NCF: no colorectal findings, SSL: sessile serrated lesion; TSA: traditional serrated adenoma

**Supplementary table 2**. **List of the 330 differentially methylated positions (DMPs)** between high-risk serrated lesions (HR-SL) and no serrated neoplasia (NSN) cfDNA pooled samples.

| | | | | | | |
|---|---|---|---|---|---|---|
| **100 hypermethylated DMPs** | | | | | | |
| cg02710296 | cg07131604 | cg06139856 | cg14970991 | cg24899334 | cg21000329 | cg18984282 |
| cg14485633 | cg25502144 | cg24917382 | cg16571209 | cg02981003 | cg10377414 | cg00901138 |
| cg15386434 | cg13560853 | cg18029503 | cg04244097 | cg14205800 | cg17864199 | cg25836915 |
| cg08222618 | cg09508496 | cg23690866 | cg27334919 | cg07770222 | cg10579706 | cg02691506 |
| cg11245681 | cg15829535 | cg24422984 | cg14977069 | cg06762332 | cg20643070 | cg02142926 |
| cg16170495 | cg02792740 | cg09911480 | cg11794430 | cg10184387 | cg17107599 | cg07165610 |
| cg11062466 | cg02873885 | cg14989243 | cg25835058 | cg22937632 | cg01927686 | cg15370054 |
| cg00050872 | cg24820663 | cg17387122 | cg07220815 | cg05322837 | cg22760004 | cg12008779 |
| cg05233899 | cg27207756 | cg16733226 | cg09533556 | cg01332534 | cg02925295 | cg11836212 |
| cg08296601 | cg15570860 | cg17847520 | cg08779649 | cg14282114 | cg24480555 | cg00541104 |
| cg16709904 | cg14663589 | cg09035930 | cg20594303 | cg20737204 | cg08177015 | |
| cg17422692 | cg13425294 | cg09929369 | cg17468267 | cg22851864 | cg01132407 | |
| cg08767686 | cg27089703 | cg20016914 | cg08369164 | cg18583021 | cg16955800 | |
| cg10482508 | cg19767205 | cg01327984 | cg02399371 | cg01307861 | cg22595420 | |
| cg05524354 | cg00965110 | cg01067216 | cg26725559 | cg00345425 | cg06574229 | |
| **230 hypomethylated DMPs** | | | | | | |
| cg10160312 | cg25755428 | cg06218861 | cg10326673 | cg14854723 | cg11424260 | cg00325531 |
| cg26266427 | cg27173819 | cg16306629 | cg08142904 | cg09389091 | cg15148984 | cg06115838 |
| cg02049405 | cg10523645 | cg10624328 | cg08661751 | cg02989453 | cg23622162 | cg08439244 |
| cg12000995 | cg27587661 | cg04902443 | cg03782861 | cg04117076 | cg06696958 | cg14281821 |
| cg20434819 | cg21820656 | cg08485684 | cg23907051 | cg21940877 | cg00635950 | cg26977859 |
| cg16967003 | cg13943068 | cg08921133 | cg23216724 | cg14777352 | cg01608493 | cg03157329 |
| cg07524919 | cg01337207 | cg15652532 | cg00035220 | cg14223966 | cg03744383 | cg24976744 |
| cg05874882 | cg03729553 | cg03369957 | cg02844899 | cg01649601 | cg19406349 | cg20433858 |
| cg19944848 | cg15165122 | cg16409883 | cg26590199 | cg21268984 | cg08560373 | cg12404279 |
| cg08269974 | cg14294859 | cg19083914 | cg10575075 | cg09251068 | cg21654286 | cg07164567 |
| cg17841765 | cg00525277 | cg00791868 | cg05018460 | cg03746345 | cg04641400 | cg04064998 |
| cg10923662 | cg07970752 | cg00182994 | cg15975750 | cg15209676 | cg22262325 | cg23304078 |
| cg23950714 | cg18373855 | cg24078577 | cg07189587 | cg15167547 | cg15684681 | cg17675992 |
| cg19754622 | cg06642012 | cg10478315 | cg25764931 | cg05785598 | cg09124484 | cg08029622 |
| cg22728830 | cg07643097 | cg05493407 | cg15176005 | cg11383474 | cg10521014 | cg03054605 |
| cg19675142 | cg20667709 | cg01952194 | cg11141652 | cg13175739 | cg20961723 | cg01652075 |
| cg05360714 | cg00872984 | cg26342559 | cg27582696 | cg04849318 | cg25620243 | cg04061506 |
| cg06157435 | cg23995446 | cg21875980 | cg18771300 | cg06736542 | cg20434529 | cg05820623 |
| cg26371957 | cg02794151 | cg20507276 | cg20067415 | cg01387720 | cg20806021 | cg20112774 |
| cg03748376 | cg27245348 | cg19567415 | cg08787791 | cg01201512 | cg01921126 | cg22093506 |
| cg07210187 | cg13526469 | cg10197405 | cg00000776 | cg14155724 | cg01035815 | cg23127434 |
| cg23670519 | cg15265085 | cg12777182 | cg15249221 | cg06704455 | cg06312072 | cg04071225 |
| cg08431882 | cg18990407 | cg04566799 | cg25482454 | cg26771832 | cg08810073 | cg15171452 |
| cg05754624 | cg05578102 | cg00079551 | cg08475528 | cg23954759 | cg19787644 | cg01293971 |
| cg16461996 | cg09506600 | cg25082212 | cg18151703 | cg20239921 | cg02881189 | cg23097878 |
| cg20839080 | cg25556122 | cg11112615 | cg24819596 | cg00799742 | cg00659250 | cg03086067 |
| cg09318283 | cg05825244 | cg01644798 | cg18961703 | cg26203738 | cg07157030 | cg26754552 |
| cg01516119 | cg01693350 | cg14703454 | cg12900080 | cg05060901 | cg26647036 | cg07248440 |
| cg07392432 | cg15399759 | cg02107357 | cg10802974 | cg02970458 | cg20276780 | cg01301660 |
| cg03012280 | cg19690306 | cg00853151 | cg14096311 | cg03553226 | cg23971638 | cg09755872 |
| cg05552010 | cg04543124 | cg20806345 | cg09959687 | cg08842287 | cg10577630 | cg16106427 |
| cg11015196 | cg08201663 | cg21263170 | cg26119671 | cg08861456 | cg22927494 | cg03660901 |
| cg23053444 | cg00956907 | cg08209240 | cg18033029 | cg07803173 | cg02181349 | |

**Supplementary table 3**. List of significantly enriched gene ontology (GO) terms in the hypermethylated and hypomethylated DMPs between high-risk serrated lesions (HR-SL) and no serrated neoplasia (NSN).

| DMP set | Ontology | GO ID | Name | Over-representation *p*-value | Differentially methylated genes overlapping with the GO term |
|---|---|---|---|---|---|
| **Hypermethylated** | BP | GO:0000495 | box H/ACA snoRNA 3'-end processing | 0.0025 | PARN |
| | BP | GO:0003199 | endocardial cushion to mesenchymal transition involved in heart valve formation | 0.0034 | TWIST1 |
| | BP | GO:0010260 | animal organ senescence | 0.0036 | COMP |
| | BP | GO:0030042 | actin filament depolymerization | 0.0046 | ADD2, MICAL3 |
| | BP | GO:0032984 | protein-containing complex disassembly | 0.0009 | MRPL28, ADD2, NCKAP5, MICAL3 |
| | BP | GO:0033128 | negative regulation of histone phosphorylation | 0.0047 | TWIST1 |
| | BP | GO:0033979 | box H/ACA snoRNA metabolic process | 0.0037 | PARN |
| | BP | GO:0034964 | box H/ACA snoRNA processing | 0.0025 | PARN |
| | BP | GO:0035359 | negative regulation of peroxisome proliferator activated receptor signaling pathway | 0.0037 | TWIST1 |
| | BP | GO:0043624 | cellular protein complex disassembly | 0.0002 | MRPL28, ADD2, NCKAP5, MICAL3 |
| | BP | GO:0051261 | protein depolymerization | 0.0009 | ADD2, NCKAP5, MICAL3 |
| | BP | GO:0090500 | endocardial cushion to mesenchymal transition | 0.0046 | TWIST1 |
| | BP | GO:0090669 | telomerase RNA stabilization | 0.0046 | PARN |
| | BP | GO:0110008 | ncRNA deadenylation | 0.0025 | PARN |
| | BP | GO:0140049 | regulation of endocardial cushion to mesenchymal transition | 0.0010 | TWIST1 |
| | BP | GO:0140051 | positive regulation of endocardial cushion to mesenchymal transition | 0.0010 | TWIST1 |
| | BP | GO:2000275 | regulation of oxidative phosphorylation uncoupler activity | 0.0045 | TWIST1 |
| | BP | GO:2000276 | negative regulation of oxidative phosphorylation uncoupler activity | 0.0023 | TWIST1 |

Supplementary Table 3. (continuation)

| DMP set | Ontology | GO ID | Name | Over-representation *p*-value | Differentially methylated genes overlapping with the GO term |
|---|---|---|---|---|---|
| **Hypermethylated** | BP | GO:2000802 | positive regulation of endocardial cushion to mesenchymal transition involved in heart valve formation | 0.0010 | TWIST1 |
| | BP | GO:2000800 | regulation of endocardial cushion to mesenchymal transition involved in heart valve formation | 0.0010 | TWIST1 |
| | CC | GO:0070033 | synaptobrevin 2-SNAP-25-syntaxin-1a-complexin II complex | 0.0050 | CPLX2 |
| | MF | GO:0004750 | ribulose-phosphate 3-epimerase activity | 0.0017 | RPEL1 |
| | MF | GO:0016232 | HNK-1 sulfotransferase activity | 0.0011 | CHST10 |
| | MF | GO:0099103 | channel activator activity | 0.0029 | LRRC26 |
| | MF | GO:0099104 | potassium channel activator activity | 0.0029 | LRRC26 |
| **Hypomethylated** | BP | GO:0003312 | pancreatic PP cell differentiation | 0.0029 | NEUROD1 |
| | BP | GO:0003326 | pancreatic A cell fate commitment | 0.0029 | NEUROD1 |
| | BP | GO:0003329 | pancreatic PP cell fate commitment | 0.0029 | NEUROD1 |
| | BP | GO:0014835 | myoblast differentiation involved in skeletal muscle regeneration | 0.0014 | WNT10B |
| | BP | GO:0045940 | positive regulation of steroid metabolic process | 0.0025 | CES1, FGF1 |
| | BP | GO:0060730 | regulation of intestinal epithelial structure maintenance | 0.0040 | NEUROD1 |
| | BP | GO:0090122 | cholesterol ester hydrolysis involved in cholesterol transport | 0.0019 | CES1 |
| | BP | GO:0090205 | positive regulation of cholesterol metabolic process | 0.0005 | CES1, FGF1 |
| | BP | GO:1903810 | L-histidine import across plasma membrane | 0.0044 | SLC7A1 |
| | MF | GO:0004771 | sterol esterase activity | 0.0020 | CES1, NCEH1 |
| | MF | GO:0004903 | growth hormone receptor activity | 0.0046 | GHR |
| | MF | GO:0046523 | S-methyl-5-thioribose-1-phosphate isomerase activity | 0.0020 | MRI1 |

BP: biological process, CC: cellular component, MF: molecular function ontologies.

**Supplementary table 4.** Primers, probes, and amplicon details for the evaluation of the DMRs in individual serum samples. The same pair of primers were used for the pre-amplification and the MS-qPCR. Probes were specific for the fully-methylated sequence. *ACTB* gene was used to normalize for DNA input.

| DMR | Primers and probe | Targeted region (GRCh37/hg19) | Length (bp) | CpG sites analyzed |
|---|---|---|---|---|
| DMR2 | Forward: GGGAGTGGGTTTAGTAATGG<br>Reverse: AAACACAATACTCAATACCTAAC<br>Probe: 5´ 6-FAM/TTATAGTTT/ZEN/CGGGTCGGGATTCGTTAG/3IABkFQ | chr12:740220-740319 | 100 | 3 |
| DMR7 | Forward: TTAGCGYGTTGGATTGATAATTG<br>Reverse: ATACCRCACCCTCCTCTC<br>Probe: 5´ 6-FAM/ACTACGCGC/ZEN/CTCATTCGACAATT/3IABkFQ | chr4:124309-124368 | 60 | 3 |
| DMR9 | Forward: TGATAGGATYGGGTTTGGGGAAG<br>Reverse: AAATCRCRCTCAACRCAAAAAAAC<br>Probe: 5´ 6-FAM/GTAGAGTTT/ZEN/GAGTATTTCGGATCGCGT/3IABkFQ | chr8:599907-600046 | 140 | 3 |
| *ACTB* | Forward: TCCCTTAAAAATTACAAAAACCACA<br>Reverse: TGGTGATGGAGGAGGTTTAG<br>Probe: 5´ 6-FAM/ACCACCACC/ZEN/CAACACACAATAACAAAAACA/3IABkFQ | chr7:5571748-5571861 | 114 | 0 |

**SUPPLEMENTARY FIGURES**

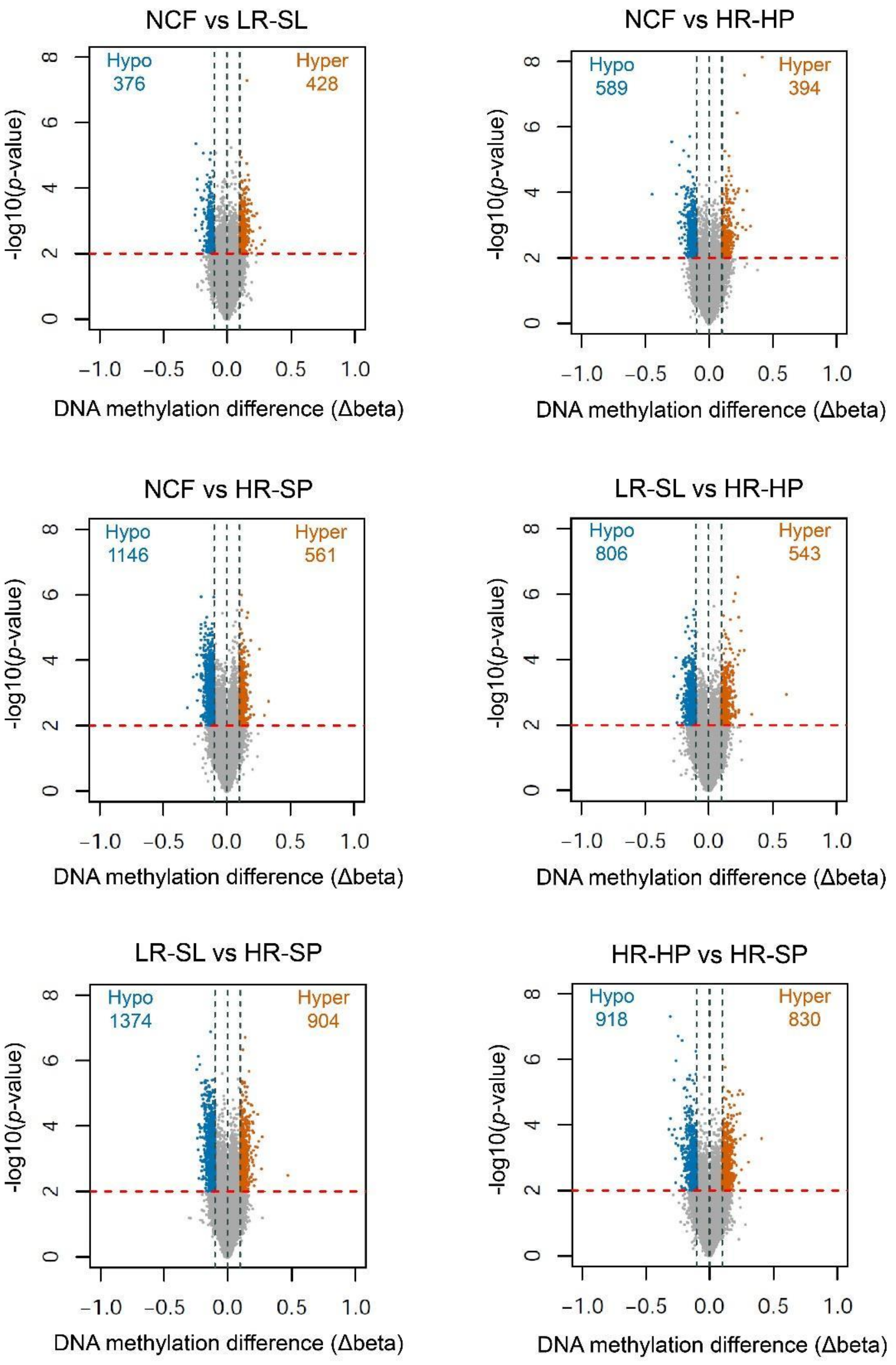


**Supplementary Figure 1. Results of the differential methylation analysis at probe level.** Volcano plots showing the -log$_{10}$(*p*-value) versus differences in methylation levels (Δbeta: obtained by subtracting the DNA methylation levels (beta-values) of the two groups involved in each analysis). Significant differentially methylated positions (DMPs) appear highlighted in blue (CpG sites with at least 10% hypomethylation in the most-severe lesion) and orange (CpG sites with at least 10% hypermethylation in the most-severe lesion), and above the red dashed line (p-value < 0.01). HR-HP: high-risk hyperplastic polyp, HR-SP: high-risk serrated polyp, LR-SL: low-risk serrated lesion, NCF: no colorectal findings.

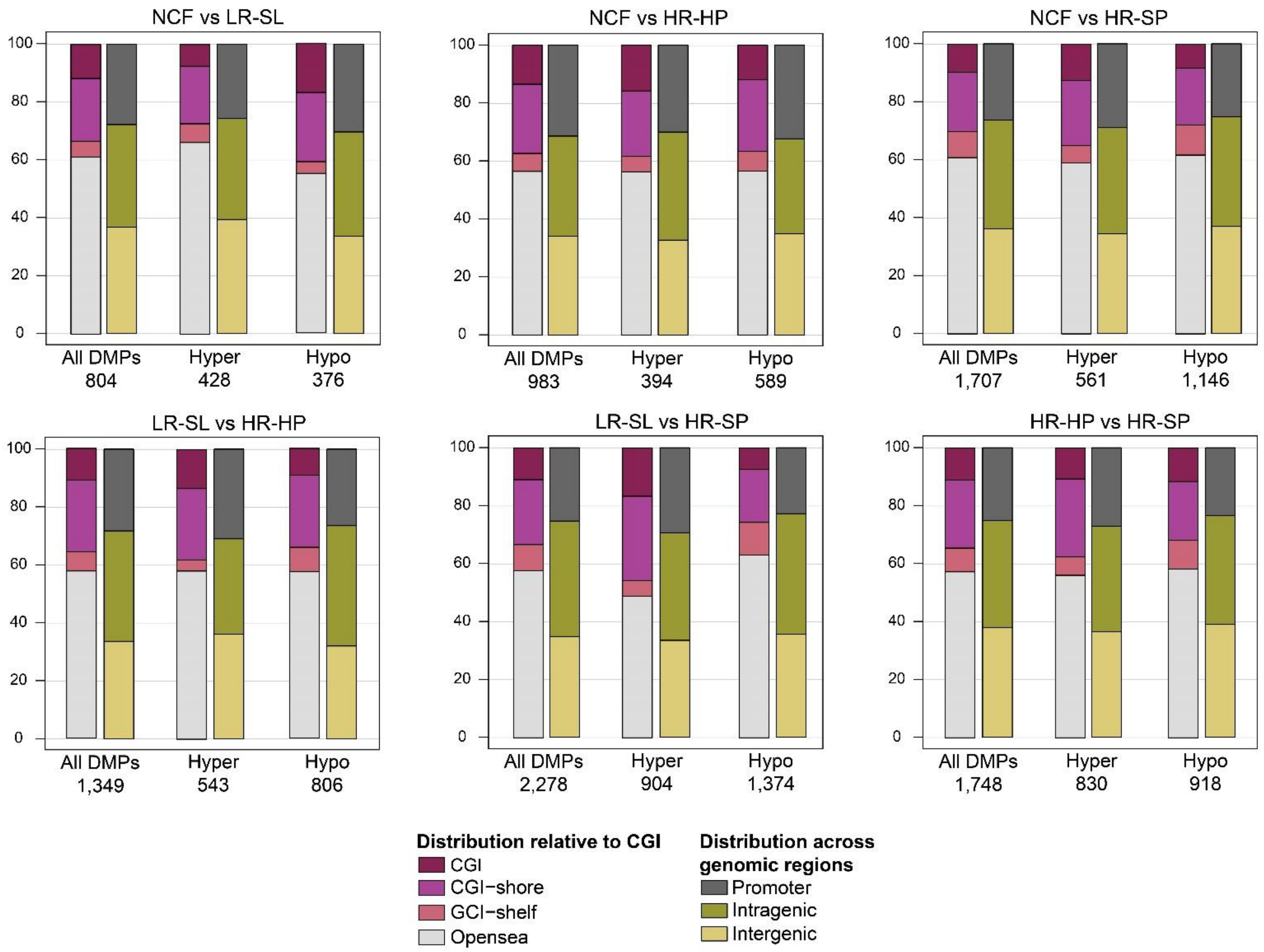


**Supplementary Figure 2**. Distribution of the DMPs obtained from all the pairwise comparisons, relative to CGI and functional genomic locations. CGI (CpG island): region of at least 200 bp with a CG content > 50% and an observed-to-expected CpG ratio ≥ 0.6; CGI-shore: sequences 2 kb flanking the CGI, CGI-shelf: sequences 2 kb flanking shore regions, opensea: sequences located outside these regions, promoter regions (5'UTR, TSS200, TSS1500, and first exons), intragenic regions (gene body and 3'UTR), and intergenic regions. TSS200, TSS1500: 200 and 200-1500 bp upstream of the transcription start site, respectively. HR-HP: high-risk hyperplastic polyp, HR-SP: high-risk serrated polyp, LR-SL: low-risk serrated lesion, NCF: no colorectal findings.

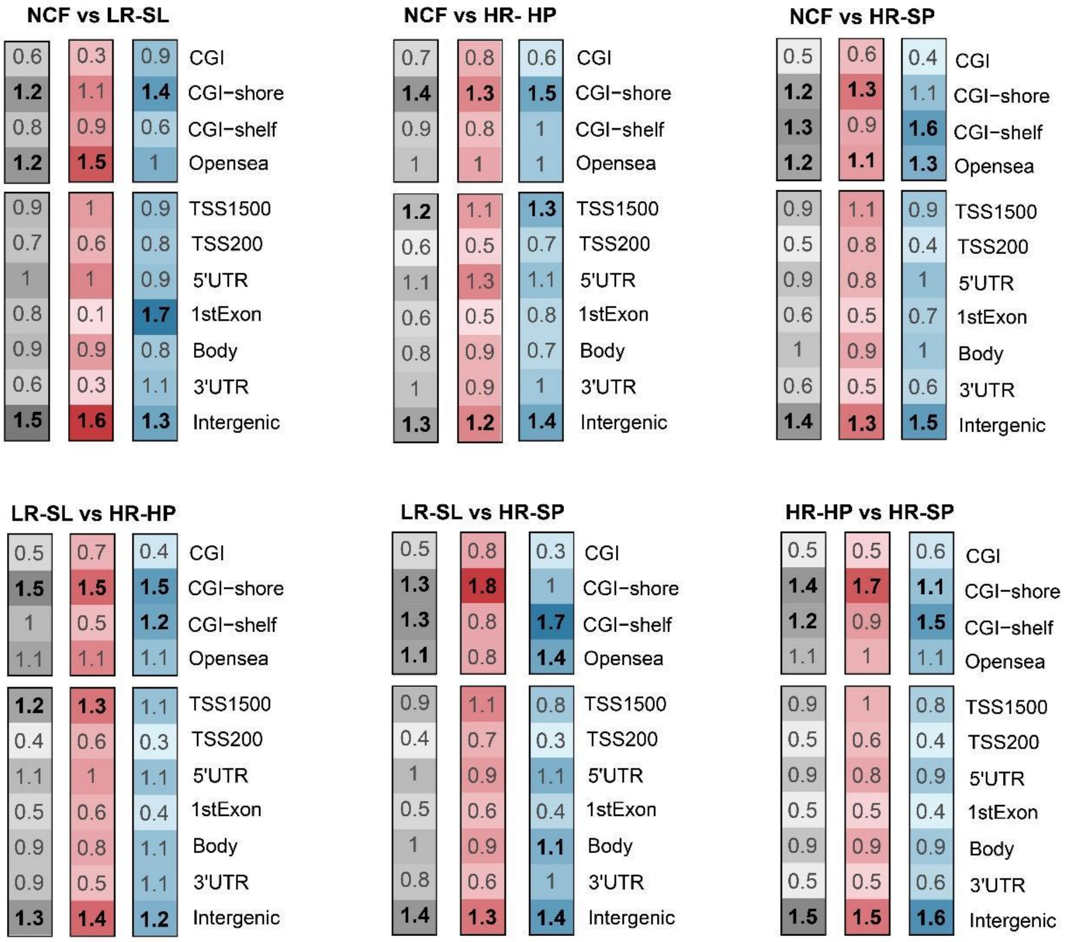


**Supplementary Figure 3.** Enrichment of DMPs obtained from all the pairwise comparisons, in relation to CGI annotation and functional genomic regions. The color scale indicates the fold enrichment of all DMPs (gray), hypermethylated (red), and hypomethylated (blue) positions. The bolded numbers indicate annotations that are enriched with respect to the distribution of probes on the MethylationEPIC array (odds-ratio > 1 and one-sided Fisher's exact test *p*-value < 0.05). CGI (CpG island): region of at least 200 bp with a CG content > 50% and an observed-to-expected CpG ratio ≥ 0.6; CGI-shore: sequences 2 kb flanking the CGI, CGI-shelf: sequences 2 kb flanking shore regions, opensea: sequences located outside these regions. TSS200, TSS1500: 200 and 200-1500 bp upstream of the transcription start site, respectively. HR-HP: high-risk hyperplastic polyp, HR-SP: high-risk serrated polyp, LR-SL: low-risk serrated lesion, NCF: no colorectal findings.

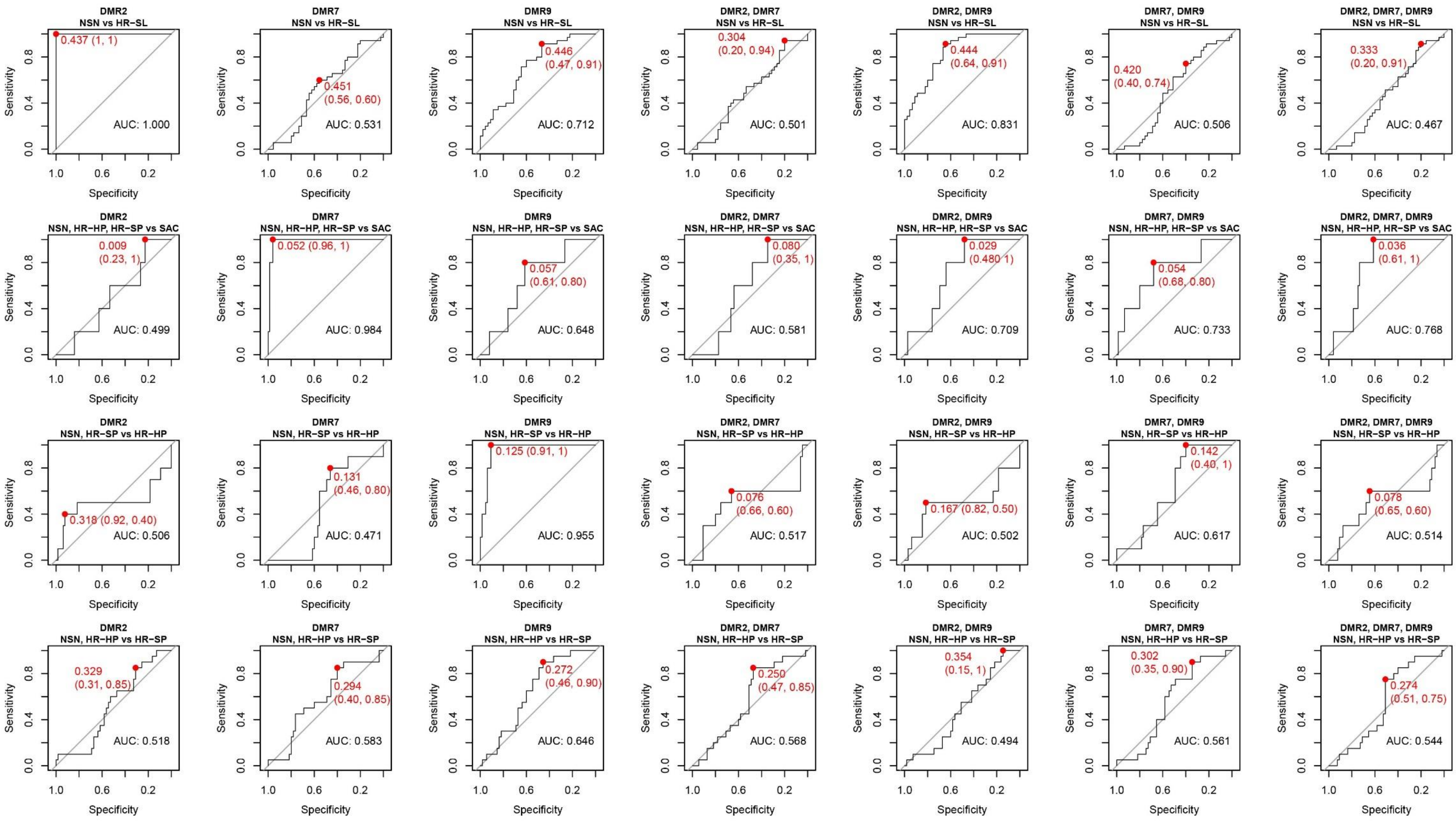


**Supplementary Figure 4.** ROC curve analysis and AUC for logistic regression models obtained with single or combinations of DMRs for the detection of HR-SL, SAC, HR-HP, or HR-SP, derived by leave-one-out cross-validation in the individual serum samples (n=80). Sensitivity and specificity values for the best cut-offs based on the Youden Index method are highlighted in red. HR-HP: high-risk hyperplastic polyp, HR-SL: high-risk serrated lesion, HR-SP: high-risk serrated polyp, NSN: no serrated neoplasia, SAC: serrated adenocarcinoma.
